# Hemoglobin levels among male agricultural workers: analyses from the Demographic and Health Surveys to investigate a marker for chronic kidney disease of uncertain etiology

**DOI:** 10.1101/2021.09.14.21263584

**Authors:** Yuzhou Lin, Siyu Heng, Shuchi Anand, Sameer K. Deshpande, Dylan S. Small

## Abstract

**Background:** Chronic kidney disease of uncertain etiology (CKDu) has been found at high frequency in several lowland agricultural areas. Whether CKDu occurs in other countries with large agricultural populations remains uncertain, primarily due to lack of systematic data on kidney function. Hemoglobin (Hgb) levels are an ancillary marker for kidney dysfunction. We estimate the causal effect of agricultural work on Hgb level in men. A causal effect may indicate the presence of CKDu.

**Methods:** We use Demographic and Health Surveys (DHS) data from seven African and Asian countries to estimate the causal effect of agricultural work on altitude-adjusted Hgb levels after adjusting for seven measured confounders. To assess potential bias due to unmeasured socioeconomic differences, we use multiple control groups that differ in non-agricultural occupation. We conduct sensitivity analyses to assess the robustness of our causal conclusions to unmeasured confounding.

**Results:** Data were available for 41,180 agricultural workers and 55,705 non-agricultural workers. On average, Hgb levels were 0.09 g/dL lower among agricultural workers compared to matched controls. Significant effects were observed in Ethiopia, India, Lesotho, and Senegal, with effects from 0.07 to 0.30 g/dL lower hemoglobin among agricultural workers.

**Conclusions:** We find evidence that men engaged in agricultural work in four of the seven countries studied have modestly lower Hgb levels compared with comparable men. Since underlying kidney disease could be a potential explanation for this finding, our data support consideration to integrating kidney function assessments within DHS surveys and other population-based surveys.

## INTRODUCTION

A kidney disease of uncertain etiology has been found to be occurring at high frequency in several lowland agricultural areas of the world including Meso-America and Sri Lanka, where it is now recognized as a leading cause of death.^1,2,3^ Whether this disease (CKDu) also occurs in other countries with large agricultural populations remains uncertain, primarily due to lack of systematic data on kidney function.

On a population-wide level, lacking data on serum creatinine assessments, hemoglobin (Hgb) levels could be an ancillary marker for presence of kidney dysfunction. Hgb is measured in several surveys that reach populations residing in low- and middle-income countries.^4,5^ Although the prevalence of frank anemia (i.e., Hgb < 13 g/dL in men and < 12 g/dL in women) is relatively low even at moderate levels of kidney dysfunction, Hgb levels start to drop early in the disease course.^6^ In an analysis from the third National Health and Nutrition Examination Survey, Astor et al. observed a decline in median Hgb levels starting at estimated glomerular filtration rate (eGFR) below 60 ml/min/1.73m^2^. Hsu et al. reported a signal starting at earlier stages of kidney dysfunction; for example, estimated Hgb were -0.2 g/dL lower among men with eGFR 60-70 compared with men with eGFR > 80 ml/min/1.73m^2^.^7^ In a meta-analysis evaluating data from over 250,000 persons with and without CKD, both the CKD and healthy cohorts demonstrated a continuous and negative relationship between eGFR and hemoglobin, starting at eGFR<60 ml/min/1.73m^2^.^8^ Finally, systemic illness including chronic kidney disease is the more likely cause of anemia among middle-aged persons, especially among men (as opposed to iron deficiency in young children and child-bearing age women).^9^ Thus, differentially low Hgb levels in our population of interest could imply the need for further investigation into kidney dysfunction as a potential cause.

The Demographic and Health Surveys (DHS) are nationally-representative household surveys primarily conducted in low- and middle-income countries that collect data on several health and sociodemographic indicators.^6^ Since Hgb levels are systematically measured in the DHS, we sought to evaluate their association with occupation, with the hypothesis that Hgb levels would be lower among men working in agriculture than among men working in other occupations after accounting for age, and nutritional and wealth indices. Such a finding is potentially indicative of an under-recognized higher prevalence of kidney dysfunction among agricultural workers, and would support rationale for integrating systematic screening for kidney disease by occupation in future DHS surveys as well as other national or regional disease surveillance systems.

## METHODS

### Study population

We obtained from the Integrated Public Use Microdata Series’ recoding of the Demographic and Health Surveys (IPUMS-DHS) the standard DHS survey data for our analysis. Standard DHS surveys, which are usually conducted in various developing countries every five years, collect comparative data on population, health, and nutrition.^10^ We picked one standard DHS survey per country using IPUMS-DHS, as IPUMS-DHS recodes some DHS variables across different surveys to make sure important data are consistent across years and surveys.^11^ We used three criteria to select country samples: (1) availability of Hgb levels; (2) availability of data on seven *a priori* identified potential confounders of hemoglobin and occupation (see Supplementary Materials Section 1 for details on the seven measured confounders); (3) samples collected in the most recent year the DHS was conducted for the country were preferred. The final selected DHS samples were from six African countries and one Asian country (surveys were conducted in different years, between 2010 and 2016), including Ethiopia in 2016, Lesotho in 2014, Namibia in 2013, Senegal in 2010, Uganda in 2016, Zimbabwe in 2015, and India in 2015.

Since most agricultural workers live in rural areas, we included only men from rural areas in our analytic sample. We exclude all men whose occupation or Hgb level were missing. A flow diagram of inclusion criteria and data pre-processing steps is available in Section 2 of the Supplementary Materials. In all, our analytic sample contained 41,180 agricultural workers and 55,705 non-agricultural workers, whose ages ranged between 15 and 64 at the time of the surveys.

### Data extraction

We extracted data on male workers’ age, body mass index, wealth index, education level, marital status, religion, occupation, degree of cluster rurality, and altitude-adjusted Hgb. DHS adjusted its measured Hgb of male workers for altitudes higher than 1,000 meters since oxygen is less available as altitude increases so effective hemoglobin count is lowered (see Supplementary Materials Section 1 for details on the adjustment).^12^ The agricultural worker category coded by IPUMS-DHS includes farmers, either self-employed or employee, as well as fishermen, foresters, breeders, and hunters.^13^ Non-agricultural workers consist of the following occupations – professional, managerial, clerical, sales, manual labor, household, domestic services, and other non-agricultural occupations – as well as those not working.

### Matching

Agricultural workers and non-agricultural workers may differ substantially in the seven measured confounders (age, body mass index, wealth index, education, marital status, religion, and degree of cluster rurality) and these confounders may affect Hgb. Consequently, a direct comparison of the average Hgb levels of the agricultural workers and nonagricultural workers may be biased. To address this potential bias, we constructed matched sets of agricultural workers and non-agricultural rural workers who are similar on the measured confounders. We then compared Hgb levels within these matched sets of comparable agricultural and nonagricultural workers. To construct the matched sets, we implemented optimal full matching^14,15^ using the R package “optmatch”^16^ for every DHS country sample. Each matched set contained either one agricultural worker and multiple non-agricultural workers, or multiple agricultural workers and one non-agricultural workers. The full matching method imposes a propensity score caliper^17^ and minimizes the rank-based Mahalanobis distance^18^ between matched male workers with similar propensity scores.

To evaluate whether the agricultural workers and controls were balanced on the measured confounders within matched sets, we calculated the standardized differences before matching and after matching. The standardized difference of a confounder before matching is the difference between the means of the confounder for the agricultural workers vs. controls (non-agricultural workers) in within group pooled standard deviation units while the standardized difference after matching is the weighted average of the difference in means within matched sets between the agricultural workers and controls in the same within group pooled standard deviation units as before matching where the weighting is by the number of agricultural workers in the matched set.^18^ The goal was to achieve adequate balances over the seven measured confounders, by making the standardized differences between the agricultural workers and controls on the seven measured confounders below 0.2 after matching.^19^ In terms of a normal distribution, 95% of the distribution is contained in a range of ±2 standard deviations, so a standardized difference of 0.2 is only 5% of the distribution, a small quantity, that can easily be removed by model based adjustments.^20,21^

### Control Groups

One potential bias for the study is that agricultural workers might have worse socioeconomic status in ways that were not fully captured by the seven measured confounders and this unmeasured socioeconomic status might affect Hgb. To assess potential bias from an unmeasured confounder, Campbell^18,22^ suggested constructing two control groups that systematically vary the unmeasured confounder and examining whether the control groups have different outcomes after controlling for measured confounders. We considered two control groups of men – (i) men who had professional, managerial, clerical, and sales occupations and (ii) men who had other non-agricultural occupations (manual labor, household, and domestic services, other and not working). Control group (i) had higher measured wealth and education (mean wealth quintile = 3.32; proportion with higher than secondary education = 31.4%) than control group (ii) (mean wealth quintile = 2.65; proportion with higher than secondary education = 9.7%) and likely higher unmeasured aspects of socioeconomic status, which could affect the outcome of interest – altitude-adjusted Hgb. Control group (ii) has higher measured wealth and education than agricultural workers (mean wealth quintile = 2.39; proportion with higher than secondary education = 4.7%) but is closer to agricultural workers than to control group (i). We call control group (i) the better-off controls and control group (ii) the worse-off controls. A detailed summary of wealth index and education level by occupation is in Section 3 of the Supplementary Materials.^18^ To check the comparability of alternative control groups, we implemented four separate full matchings over the seven measured confounders for the following four comparisons: agricultural vs. all controls, agricultural vs. better-off controls, agricultural vs. worse-off controls, and worse-off vs. better-off.

### Permutation Inference

To estimate the treatment effect of agricultural occupation on the outcome, altitude-adjusted Hgb levels, we conducted permutation inferences on the matched DHS samples. The tested null hypothesis was that agricultural occupation would have no effect on men’s Hgb levels, and the alternative hypothesis was that agricultural occupation would lower men’s Hgb levels. To further reduce the bias for estimating the treatment effect due to measured confounding, we combined the techniques of permutation test and covariance adjustment – regressing men’s Hgb levels on the measured confounders and then conducting permutation tests on the residuals from matched sets.^20,21^ Specifically, we used Huber’s m-statistics for matched sets to calculate both the lower bounds and the upper bounds for the one-sided p-values^23^ (using the “senfm” function with default parameters in the R package “sensitivityfull”^24^) and the 95% upper confidence bound under the additive treatment effect model. We first conducted permutation tests for the overall effect on the whole matched sample. Then we tested country-specific effects for the seven matched DHS samples individually. We adjusted upper-bounded p-values from testing for the whole matched sample and the seven matched DHS samples to control the false discovery rate using the Benjamini-Hochberg approach.^25,26^ To check if the alternative control groups were comparable (to test for bias from unmeasured aspects of socioeconomic status as described above), we estimated the treatment effects of occupation on altitude-adjusted Hgb levels in each of the four comparisons (agricultural vs. all controls, agricultural vs. better-off controls, agricultural vs. worse-off controls, and worse-off vs. better-off.). Testing for the four comparisons followed an ordered hypothesis testing procedure which controls the familywise error rate for multiple testing at level .05.^27^

### Sensitivity Analysis

The permutation inference results of our primary analysis assume there are no unmeasured confounders. We assessed how sensitive the results of our primary analysis were to violations of the no unmeasured confounding assumption. Using Huber’s m-statistics, we conducted sensitivity analyses first on the whole sample and then on each DHS country sample that was identified with statistically significant effects (p-value < 0.05) using the approach introduced by Rosenbaum.^19^ In the sensitivity analysis, we consider different possible values of the sensitivity parameter Γ which is, for two participants with the same measured confounders, the maximum the odds ratio for being an agricultural worker could be for one participant compared to the other because of unmeasured confounding variables. For example, if the unmeasured confounding consisted of HIV status, Γ would be the odds ratio that a person with HIV would be an agricultural worker compared to a person with the same measured confounders (i.e., age, BMI, education, marital status, religion, and wealth quintile) who does not have HIV. If Γ = 1, then there is no unmeasured confounding while the more Γ departs from 1, the more unmeasured confounding there is. We cannot know Γ since it is determined by unobserved variables, but we can consider different possible Γ, compute both a lower bound and an upper bound for the p-value that tests whether there is a treatment effect if that Γ were true, and continue until the p-value upper bound is greater than 0.05 to determine the sensitivity value, the maximum amount of unmeasured confounding there could be and still obtain a significant effect of treatment. The larger the sensitivity value is, the more robust the study’s conclusions are to unmeasured confounding. We performed the sensitivity analysis using the R package “sensitivityfull”.

## RESULTS

### Descriptive Statistics

**Table 1** summarizes the seven measured confounders among agricultural and non-agricultural workers in rural areas. In our study population, agricultural workers tended to be older and have slightly lower body mass index than non-agricultural workers. They were more likely to be married, have lower educational attainment, and fall in the bottom two wealth quintiles than non-agricultural workers. **Figure 1** displays the altitude-adjusted Hgb levels for both the agricultural and all control workers across seven DHS country samples before full matching. Altitude-adjusted Hgb levels varied significantly by country. The median altitude-adjusted Hgb levels across all samples’ treated and all control groups were between 13.5 and 15.1 g/dL.

**TABLE 1.** Mean of the seven measured confounders. Except for age, BMI, and degree of cluster rurality, we report the count (percentage) of agricultural and non-agricultural workers in each category. For age, BMI, and degree of cluster rurality, we report the mean (standard deviation).

| Correlate | Agricultural Workers<br>(n = 41180) | Non-agricultural Workers<br>(n = 55705) |
| --- | --- | --- |
| Age (years) | 34.32 (11.04) | 29.20 (10.99) |
| BMI (kg/m<sup>2</sup>) | 20.99 (3.31) | 21.20 (3.62) |
| Degree of Cluster Rurality (%) | 61.74 (23.75) | 28.28 (23.01) |
| Sample (n(%)) |  |  |
| Ethiopia 2016 | 5829 (14.2) | 2320 ( 4.2) |
| India 2015 | 29706 (72.1) | 44542 (80.0) |
| Lesotho 2014 | 605 ( 1.5) | 550 ( 1.0) |
| Namibia 2013 | 367 ( 0.9) | 1727 ( 3.1) |
| Senegal 2010 | 1327 ( 3.2) | 1187 ( 2.1) |
| Uganda 2016 | 2129 ( 5.2) | 1908 ( 3.4) |
| Zimbabwe 2015 | 1217 ( 3.0) | 3471 ( 6.2) |
| Education (n(%)) |  |  |
| None | 10374 (25.2) | 6402 (11.5) |
| Primary school | 11087 (26.9) | 9469 (17.0) |
| Secondary school | 17778 (43.2) | 32180 (57.8) |
| Higher education | 1941 ( 4.7) | 7654 (13.7) |
| Marital Status (n(%)) |  |  |
| Never married | 9562 (23.2) | 25170 (45.2) |
| Married/partnered | 30578 (74.3) | 29522 (53.0) |
| Formerly married | 1040 ( 2.5) | 1013 ( 1.8) |
| Religion (n(%)) |  |  |
| No religion | 403 ( 1.0) | 897 ( 1.6) |
| Muslim | 6171 (15.0) | 8547 (15.3) |
| Christian | 9653 (23.4) | 10063 (18.1) |
| Buddhist | 563 ( 1.4) | 644 ( 1.2) |
| Hindu | 23151 (56.2) | 33609 (60.3) |
| Jewish | 1 ( 0.0) | 5 ( 0.0) |
| Traditional | 107 ( 0.3) | 107 ( 0.2) |
| Other (Specified) | 581 ( 1.4) | 1114 ( 2.0) |
| Other | 550 ( 1.3) | 719 ( 1.3) |
| Wealth quintile (n(%)) |  |  |
| 1<sup>st</sup> quintile | 12025 (29.2) | 11245 (20.2) |
| 2<sup>nd</sup> quintile | 11880 (28.8) | 13519 (24.3) |
| 3<sup>rd</sup> quintile | 9176 (22.3) | 13875 (24.9) |
| 4<sup>th</sup> quintile | 5598 (13.6) | 10716 (19.2) |
| 5<sup>th</sup> quintile | 2501 ( 6.1) | 6350 (11.4) |

**FIGURE. 1:**
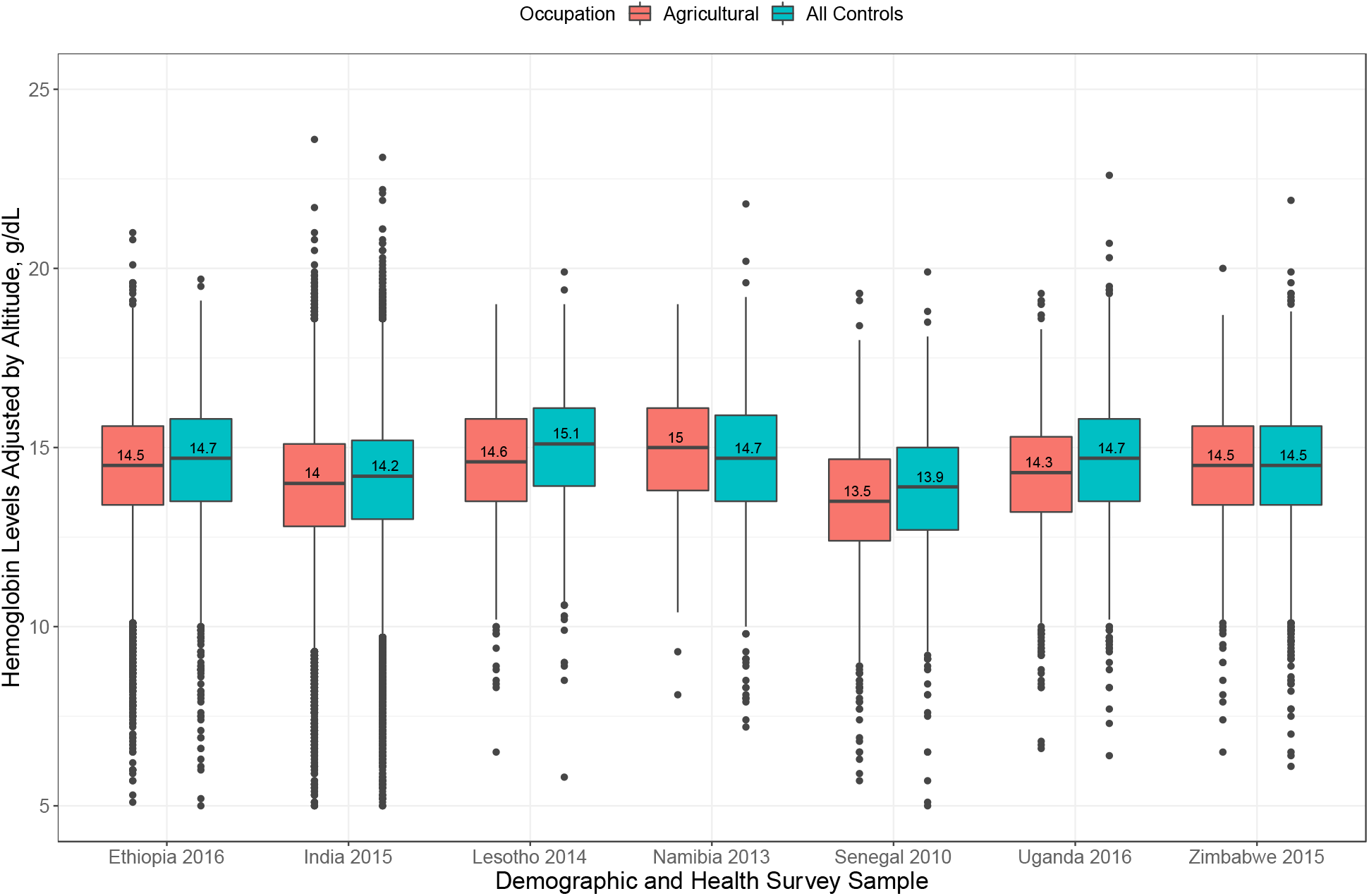
Boxplots for altitude-adjusted Hgb (g/dL) by DHS samples.

### Result of Matching

We calculated the standardized differences before and after matching for agricultural workers vs. all controls from seven DHS country samples respectively (Section 4 of the Supplementary Materials). Before matching, the absolute values of the standardized differences for most confounders were larger than 0.2 for most country samples, which indicates that the two groups were not adequately balanced. After matching, all the absolute values of the standardized differences were reduced below 0.2, suggesting adequate balances on the seven confounders was achieved. Matching for the comparisons of the agricultural workers vs. each control group and the comparison of control groups also achieved adequate balances.

### Effect of Agricultural Work

Main results for estimating the treatment effects of agricultural occupation on altitude-adjusted Hgb are in **Table 2**. We calculated the 95% confidence intervals and p-values for testing the null hypothesis of no treatment effect for all country samples and after matching agricultural workers with all controls. From **Table 2**, in our whole dataset after adjusting for potential confounding, we estimated the average effect of agricultural work on adjusted hemoglobin levels is -0.09 g/dL with a 95% upper confidence bound (UCB) of -0.06 g/dL. Among the seven countries’ samples, four countries had statistically significant (false-discovery-rate-adjusted p-value < 0.05) effects – Ethiopia (−0.13 g/dL; 95% UCB: -0.04 g/dL), India (−0.07 g/dL; 95% UCB: -0.04 g/dL), Lesotho (−0.30 g/dL; 95% UCB: -0.08 g/dL), and Senegal (−0.16 g/dL; 95% UCB: -0.02 g/dL).

**TABLE 2.** Estimated effects of agricultural work on altitude-adjusted Hgb (g/dL) after matching agricultural workers with all controls by DHS samples. Point estimate (PE) of effect and 95% upper confidence bound (UCB). Single asterisks if the false discovery rate adjusted p-value upper bound is <0.05 and double asterisk if the false discovery rate adjusted p-value upper bound is <0.001.

| DHS Sample | Number of Agricultural Workers | Number of All Control Workers | Hemoglobin Difference PE (95% UCB) |
| --- | --- | --- | --- |
| Whole DHS Sample | 41180 | 55705 | -0.09 (-0.06) ** |
| Ethiopia 2016 | 5829 | 2320 | -0.13 (-0.04) * |
| India 2015 | 29706 | 44542 | -0.07 (-0.04) ** |
| Lesotho 2014 | 605 | 550 | -0.30 (-0.08) * |
| Namibia 2013 | 367 | 1727 | 0.06 (0.28) |
| Senegal 2010 | 1327 | 1187 | -0.16 (-0.02) * |
| Uganda 2016 | 2129 | 1908 | -0.07 (0.04) |
| Zimbabwe 2015 | 1220 | 3482 | -0.04 (0.06) |

### Test for Hidden Bias

**Table 3** shows the estimated treatment effects of agricultural occupations vs. occupations in multiple control groups and better-off occupations vs. worse-off occupations on male workers’ adjusted Hgb levels. Overall, agricultural workers had lower altitude-adjusted Hgb levels than each control group. All effects were significant using the ordered testing procedure. Agricultural workers had a more negative estimated average effect vs. better-off controls (−0.16 g/dL) than worse-off controls (−0.09 g/dL). The worse-off controls had significantly lower altitude-adjusted Hgb than the better-off controls (−0.08 g/dL; 95% UCB: -0.04 g/dL).

**TABLE 3.**
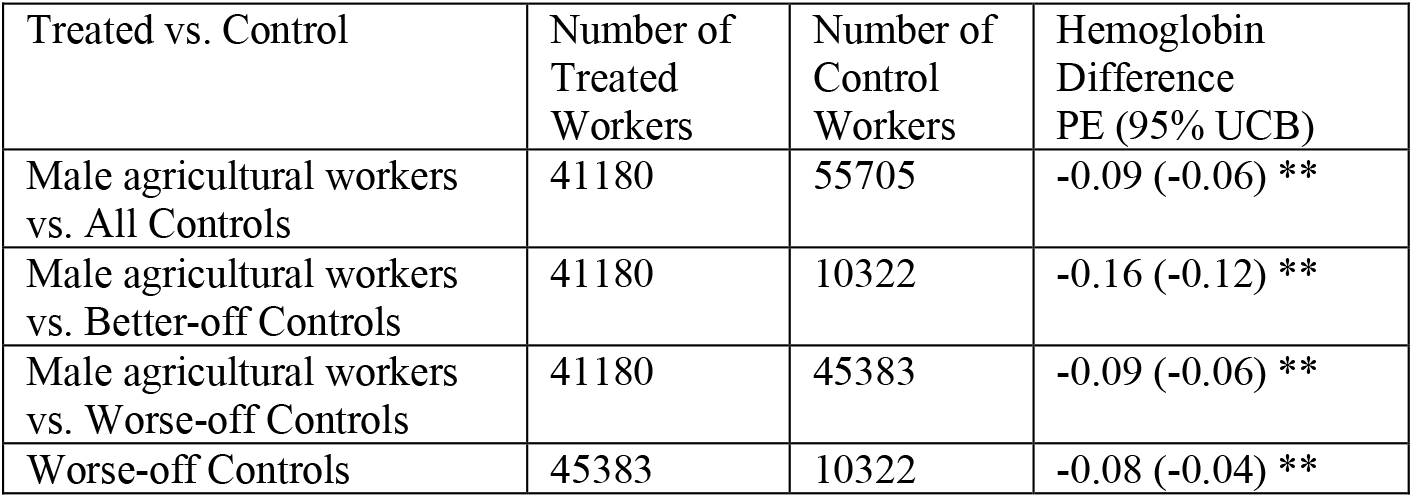
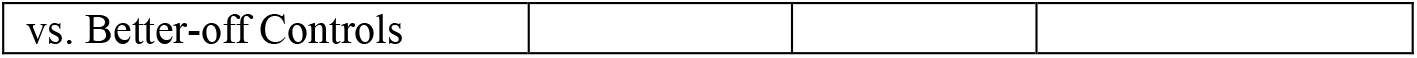
Estimated effects of agricultural work on altitude-adjusted Hgb (g/dL) after matching agricultural workers with all controls by treated and multiple controls for the whole DHS sample. Point estimate (PE) of effect and 95% % upper confidence bound (UCB). Single asterisk if the adjusted p-value upper bound (from ordered testing) < 0.05, and two asterisks if the adjusted p-value upper bound < 0.001.

### Result of Sensitivity Analysis

From **Table 2**, the whole sample consisted of all seven selected DHS samples and samples including Ethiopia, India, Lesotho, Senegal, and Uganda were respectively identified with statistically significant treatment effects. The result of sensitivity analysis for these samples is in Section 5 of the Supplementary Materials. For the whole sample, the sensitivity value of Γ to reach p-value upper bound 0.05 was 1.08. This means that if there were an unmeasured confounder that increases the odds of being an agricultural worker by 7%, we still have evidence (p-value upper bound < 0.05) that being an agricultural worker causes a reduction in Hgb. For the four country samples, the sensitivity values Γ to reach p-value upper bound 0.05 were between 1.03 and 1.10. Lesotho had the highest sensitivity value (1.10), and Senegal had the lowest sensitivity value (1.03).

## DISCUSSION

In this study of rural men participating in DHS surveys in six African and one Asian country, we found a consistent and modest effect of agricultural work on Hgb, indicating that agricultural workers have lower Hgb levels than other men residing in rural areas. This effect held up when a quasi-experimental device – multiple control groups – was used to examine concerns about unmeasured confounding. Since kidney dysfunction may be one potential explanation for our finding, we posit that our data provide rationale for systematic surveys of kidney function by occupation, especially in countries with large proportions of populations engaged in agricultural work.

Our observed effect size is modest (Hgb lower by ∼ 0.1 g/dL among agricultural workers versus other rural men), but it is concordant with effect sizes described in prior studies among persons with mild kidney dysfunction.^6,7,8,9^ We expect participation in DHS to be subject to survivor bias, and thus few, if any, persons who develop advanced kidney disease and substantial anemia, are likely present in our analytic cohort. Furthermore, our findings were robust to consideration of multiple control groups of occupations that tend to have higher and lower socioeconomic status, and a modest amount of unmeasured confounding.

Hgb level is an admittedly imperfect surrogate marker of kidney dysfunction. That said, few population-based data exist on kidney function and incidence of kidney dysfunction in agricultural communities. Even among the best described hotspots of “CKDu”, systematic surveys of prevalence and incidence of kidney dysfunction are lacking. Without better mapping of affected regions and populations, identification – and prevention – of risk factors for development of kidney dysfunction in marginalized populations is unlikely. The International Society of Nephrology has put forth a simple minimal data set inclusive of serum creatinine and urine dipsticks to assist with integration of kidney function in population surveys.^28^

Laudable investigative efforts have focused on the best-described hotspots of CKDu in Sri Lanka and in Mesoamerican countries.^2,3,29,30,31^ However, the cause of CKDu remains unclear for many reasons, including due to the potentially long lag between exposure and disease, lack of advanced research infrastructure, and political unrest. If a broader link between agricultural work and CKDu is confirmed – as has been suggested as plausible by at least two studies from the US as well^32,33^ – this could increase the desirability of in-depth investigations into cause. Our analysis is an attempt to evaluate the possibility of this broader link, using available data, and supports further direct investigations of kidney function by occupation.

The limitations of our study include limited data on confounders of the relationship between occupation and hemoglobin including water quality or HIV status. Either the DHS did not measure these factors or there were too many missing values for the sample surveys we selected. In addition, since the DHS did not separately categorize 16,798 skilled and unskilled manual laborers, we were not able to put them into different alternative control groups for the four comparisons. However the two primary confounders we expected to influence the relationship between occupation and hemoglobin levels were nutrition and socioeconomic status. We accounted for these major confounders using body mass index, marital status, and religion (which can influence diet) for the former, and wealth index, educational level, and degree of rurality for the latter. Furthermore, our matching procedure resulted in near perfectly balanced distribution of these confounders among the agricultural workers and their non-agricultural worker counterparts.

We recommend that DHS and other population-based surveys add measurements about agricultural workers’ kidney function assessments in their future standard surveys for African and Asian countries. Further studies are warranted investigating the link between agricultural work and kidney dysfunction in the four countries – Ethiopia, India, Lesotho, and Senegal – where we did find evidence of lower Hgb among agricultural workers.

## Data Availability

We obtained from the Integrated Public Use Microdata Series' recoding of the Demographic and Health Surveys (IPUMS-DHS) the standard DHS survey data for our analysis.

## ACKNOWLEDGEMENT

None declared.

## FUNDING

This research was supported in part by the University of Wisconsin – Madison, Office of the Vice Chancellor for Research and Graduate Education with funds from the Wisconsin Alumni Research Foundation. Dr. Shuchi Anand was supported by R01DK127138.

## COMPETING INTERESTS

None declared.

## Notes

### Competing Interest Statement

The authors have declared no competing interest.

### Author Declarations

No human was involved in this study.

