## Supplementary Materials for "Hemoglobin levels among male agricultural workers: analyses from the Demographic and Health Surveys to investigate a marker for chronic kidney disease of uncertain etiology"

##### Section 1. Explanation on the IPUMS-DHS Variables

**Seven *a priori* identified correlates of hemoglobin levels:**

1. Age: the participant's age at the time of the survey.
2. Body Mass Index: a 4-digit numeric variable with two implied decimal places representing a BMI for male household members.
3. Wealth Index: the relative wealth of the participant's household, divided into quintiles from the poorest (1) to the richest (5).
4. Religion: the participant 's religion. IPUMS-DHS uses a 4-digit composite coding system to fit these diverse categories into a single variable without losing information. [No religion (0); Muslim (1000); Christian (2000); Buddhist/neo-Buddhist (3000); Hindu (4000); Jewish (5000); Traditional/spiritual/animist (6000); Other (specified) (7000); Other (9000)]
5. Education Level: the highest level of school the respondent attended. This is a standardized variable reporting level of education in four categories: No education (0), Primary (1), Secondary (2), Higher (3).
6. Marital Status: the participant 's marital status at the time of the survey. [Never Married (00); Married or living together (10); Formerly in union (20)]
7. Degree of Cluster Rurality: the proportion of agricultural male workers out of all male workers in the DHS cluster where the participant lived in. We created this variable ourselves.

**Outcome:**

1. Hemoglobin Level Adjusted by Altitude: For adult men household members, HWMHEMOLEVELALT (HB56) reports the level of hemoglobin in the blood, in terms of grams per deciliter (g/dl), based on testing of blood drawn by DHS personnel, and adjusted for altitude. Hemoglobin levels are tested to determine the presence and severity of anemia. HWMHEMOLEVELALT is a 3-digit variable with 1 implied decimal place.

The adjustment followed the formulas: (1) $adjust = -0.032 \times alt + 0.022 \times alt^2$ (2) $adjusted\ Hgb = Hgb - adjust,$ if $adjust > 0$. *adjust* is the amount of the adjustment, *alt* is altitude in 1,000 feet, and *Hgb* is the measured Hgb in grams per deciliter.

**Exposure of Interest:**

1. Participant's Occupation: the occupation of the male respondent in somewhat standardized categories. The categories included, and the degree of detail within broad categories (e.g., self-employed versus employee in agriculture, or simply agriculture), vary across samples. IPUMS-DHS employs composite coding to maximize comparability across samples (using the first digit of the codes to indicate broad job categories) without loss of detailed information (preserved in the second digit). The category "agriculture" includes fishermen, foresters, breeders, and hunters as well as farmers.

### Section 2. Flow Diagram for Data Pre-processing Steps

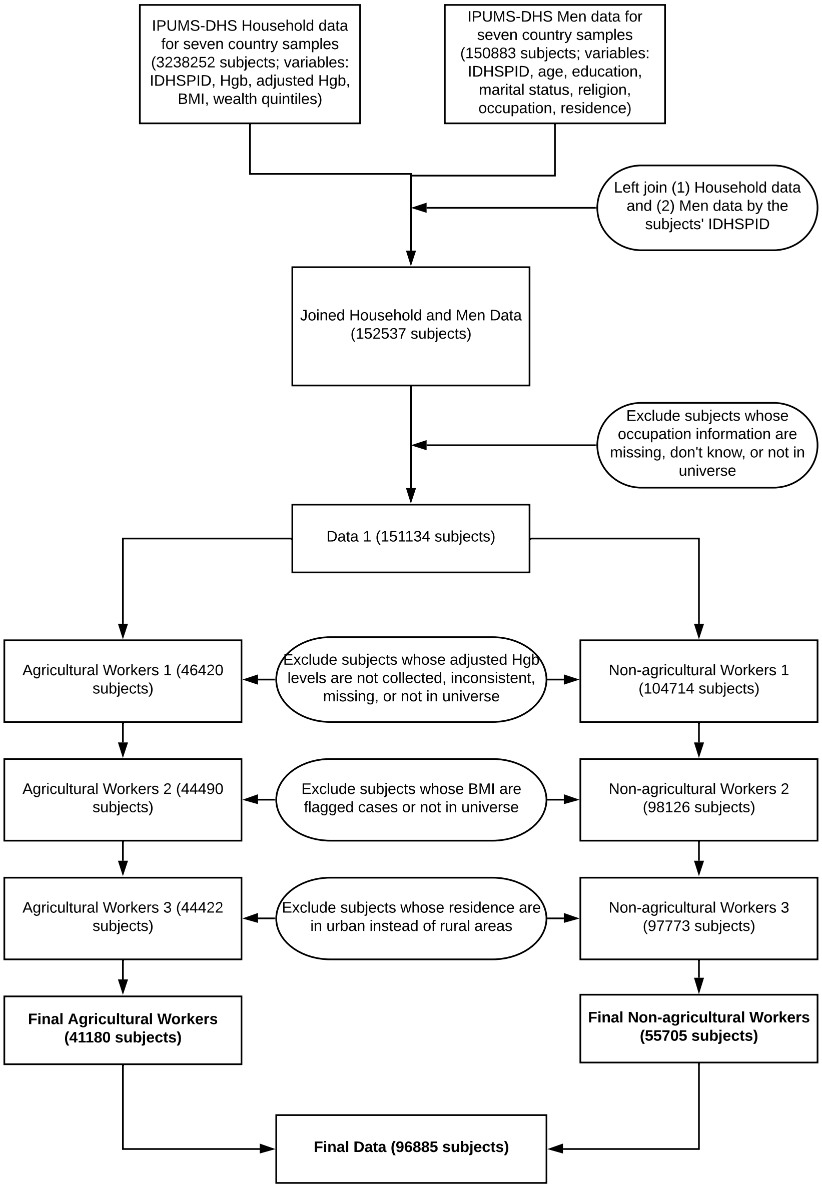

#### Section 3. Wealth Index and Education Level by Male Workers' Occupations, Treated, and Multiple Controls

| **Male Workers' Occupation** | **Size** | **Wealth Index**<br>**n (%)**<br>**1st Quintile - 1**<br>**2nd Quintile - 2**<br>**3rd Quintile - 3**<br>**4th Quintile - 4**<br>**5th Quintile - 5** | **Education Level**<br>**n (%)**<br>**no education - 0**<br>**primary education - 1**<br>**secondary education - 2**<br>**higher education - 3** |
|---|---|---|---|
| Not Currently Working | 19107 | 3806 ( 19.9)<br>4780 ( 25.0)<br>4829 ( 25.3)<br>3590 ( 18.8)<br>2102 ( 11.0) | 1285 ( 6.7)<br>2159 ( 11.3)<br>12738 ( 66.7)<br>2925 ( 15.3) |
| Professional, Technical, or Managerial | 3841 | 269 ( 7.0)<br>467 ( 12.2)<br>834 ( 21.7)<br>1164 ( 30.3)<br>1107 ( 28.8) | 85 ( 2.2)<br>179 ( 4.7)<br>1338 ( 34.8)<br>2239 ( 58.3) |
| Clerical | 1092 | 68 ( 6.2)<br>175 ( 16.0)<br>299 ( 27.4)<br>302 ( 27.7)<br>248 ( 22.7) | 30 ( 2.7)<br>79 ( 7.2)<br>648 ( 59.3)<br>335 ( 30.7) |
| Sales | 5389 | 734 ( 13.6)<br>1124 ( 20.9)<br>1387 ( 25.7)<br>1274 ( 23.6)<br>870 ( 16.1) | 684 ( 12.7)<br>941 ( 17.5)<br>3101 ( 57.5)<br>663 ( 12.3) |
| Agricultural | 37664 | 10844 ( 28.8)<br>10793 ( 28.7)<br>8408 ( 22.3)<br>5229 ( 13.9)<br>2390 ( 6.3) | 9329 ( 24.8)<br>9794 ( 26.0)<br>16627 ( 44.1)<br>1914 ( 5.1) |
| Agricultural, Self Employed | 2891 | 1076 ( 37.2)<br>878 ( 30.4)<br>564 ( 19.5)<br>270 ( 9.3)<br>103 ( 3.6) | 953 ( 33.0)<br>972 ( 33.6)<br>943 ( 32.6)<br>23 ( 0.8) |
| Agricultural, Employee | 625 | 105 ( 16.8)<br>209 ( 33.4)<br>204 ( 32.6)<br>99 ( 15.8)<br>8 ( 1.3) | 92 ( 14.7)<br>321 ( 51.4)<br>208 ( 33.3)<br>4 ( 0.6) |
| Household, Domestic, and Services | 4189 | 633 ( 15.1)<br>939 ( 22.4)<br>1137 ( 27.1)<br>945 ( 22.6)<br>535 ( 12.8) | 441 ( 10.5)<br>571 ( 13.6)<br>2542 ( 60.7)<br>635 ( 15.2) |
| Household and Domestic | 200 | 39 ( 19.5)<br>52 ( 26.0)<br>58 ( 29.0)<br>37 ( 18.5)<br>14 ( 7.0) | 11 ( 5.5)<br>92 ( 46.0)<br>95 ( 47.5)<br>2 ( 1.0) |

| | | | |
|---|---|---|---|
| Services | 1210 | 282 ( 23.3)<br>322 ( 26.6)<br>328 ( 27.1)<br>218 ( 18.0)<br>60 ( 5.0) | 136 ( 11.2)<br>566 ( 46.8)<br>486 ( 40.2)<br>22 ( 1.8) |
| Skilled and Unskilled Manual | 16798 | 4488 ( 26.7)<br>4736 ( 28.2)<br>3999 ( 23.8)<br>2463 ( 14.7)<br>1112 ( 6.6) | 3110 ( 18.5)<br>3150 ( 18.8)<br>9854 ( 58.7)<br>684 ( 4.1) |
| Skilled Manual | 2596 | 559 ( 21.5)<br>635 ( 24.5)<br>681 ( 26.2)<br>485 ( 18.7)<br>236 ( 9.1) | 327 ( 12.6)<br>1138 ( 43.8)<br>1021 ( 39.3)<br>110 ( 4.2) |
| Unskilled Manual | 722 | 183 ( 25.3)<br>176 ( 24.4)<br>192 ( 26.6)<br>138 ( 19.1)<br>33 ( 4.6) | 179 ( 24.8)<br>326 ( 45.2)<br>205 ( 28.4)<br>12 ( 1.7) |
| Other | 561 | 184 ( 32.8)<br>113 ( 20.1)<br>131 ( 23.4)<br>100 ( 17.8)<br>33 ( 5.9) | 114 ( 20.3)<br>268 ( 47.8)<br>152 ( 27.1)<br>27 ( 4.8) |

| **Agricultural workers and Multiple Controls** | **Size** | **Wealth Index**<br>**n (%)**<br>**1st Quintile - 1**<br>**2nd Quintile - 2**<br>**3rd Quintile - 3**<br>**4th Quintile - 4**<br>**5th Quintile - 5** | **Education Level**<br>**n (%)**<br>**no education - 0**<br>**primary education - 1**<br>**secondary education - 2**<br>**higher education - 3** |
|---|---|---|---|
| Agricultural Workers | 41180 | 12025 (29.2)<br>11880 (28.8)<br>9176 (22.3)<br>5598 (13.6)<br>2501 ( 6.1) | 10374 (25.2)<br>11087 (26.9)<br>17778 (43.2)<br>1941 ( 4.7) |
| All Controls | 55705 | 11245 (20.2)<br>13519 (24.3)<br>13875 (24.9)<br>10716 (19.2)<br>6350 (11.4) | 6402 (11.5)<br>9469 (17.0)<br>32180 (57.8)<br>7654 (13.7) |
| Better-off Controls | 10322 | 1071 (10.4)<br>1766 (17.1)<br>2520 (24.4)<br>2740 (26.5)<br>2225 (21.6) | 799 ( 7.7)<br>1199 (11.6)<br>5087 (49.3)<br>3237 (31.4) |
| Worse-off Controls | 45383 | 10174 (22.4)<br>11753 (25.9)<br>11355 (25.0)<br>7976 (17.6)<br>4125 ( 9.1) | 5603 (12.3)<br>8270 (18.2)<br>27093 (59.7)<br>4417 ( 9.7) |

#### Section 4. Comparison of mean baseline correlates for (1) agricultural male workers vs all controls by DHS samples, (2) agricultural male workers vs alternative controls, and (3) better-off control vs. worse-off control, before and after matching

| Whole DHS Sample | | | | | | |
|---|---|---|---|---|---|---|
| | Agricultural Male Workers | | All Controls | | Standardized Difference | |
| **Covariates** | Mean Before Matching | Mean After Matching | Mean Before Matching | Mean After Matching | Std Before Matching | Std After Matching |
| Age | 34.32 | 32.36 | 29.20 | 32.46 | 0.47 | -0.01 |
| Body Mass Index | 20.99 | 21.25 | 21.20 | 21.28 | -0.06 | -0.01 |
| Wealth Index | 2.38 | 2.56 | 2.77 | 2.56 | -0.31 | 0.00 |
| Education Level | 1.27 | 1.52 | 1.74 | 1.52 | -0.54 | 0.00 |
| Currently Married or Not | 0.74 | 0.67 | 0.53 | 0.67 | 0.45 | -0.01 |
| Christian or Not | 0.23 | 0.20 | 0.18 | 0.20 | 0.13 | 0.00 |
| Muslim or Not | 0.15 | 0.15 | 0.15 | 0.15 | -0.01 | 0.00 |
| Hindu or Not | 0.56 | 0.59 | 0.60 | 0.59 | -0.08 | 0.00 |
| No Religion or Not | 0.01 | 0.02 | 0.02 | 0.02 | -0.06 | -0.01 |
| Degree of Cluster Rurality | 0.62 | 0.46 | 0.28 | 0.46 | 1.43 | 0.01 |

| Ethiopia 2016 | | | | | | |
|---|---|---|---|---|---|---|
| Age | 32.87 | 28.84 | 26.76 | 28.63 | 0.55 | 0.02 |
| Body Mass Index | 19.27 | 19.12 | 18.93 | 19.14 | 0.14 | -0.01 |
| Wealth Index | 2.51 | 2.62 | 2.60 | 2.63 | -0.07 | -0.01 |
| Education Level | 0.69 | 0.97 | 1.16 | 1.01 | -0.06 | -0.05 |
| Currently Married or Not | 0.70 | 0.51 | 0.45 | 0.52 | 0.53 | 0.00 |
| Christian or Not | 0.60 | 0.60 | 0.54 | 0.60 | 0.12 | 0.01 |
| Muslim or Not | 0.39 | 0.38 | 0.48 | 0.38 | -0.12 | 0.01 |
| Degree of Cluster Rurality | 0.79 | 0.68 | 0.52 | 0.65 | 1.23 | 0.12 |

| India 2015 | | | | | | |
|---|---|---|---|---|---|---|
| Age | 35.28 | 33.14 | 29.31 | 33.16 | 0.56 | 0.00 |
| Body Mass Index | 21.40 | 21.52 | 21.37 | 21.54 | 0.01 | -0.01 |
| Wealth Index | 2.39 | 2.58 | 2.82 | 2.58 | -0.34 | 0.00 |
| Education Level | 1.42 | 1.62 | 1.82 | 1.62 | -0.47 | 0.00 |
| Currently Married or Not | 0.79 | 0.71 | 0.55 | 0.71 | 0.53 | -0.01 |
| Christian or Not | 0.08 | 0.07 | 0.06 | 0.06 | 0.10 | 0.00 |
| Muslim or Not | 0.08 | 0.11 | 0.14 | 0.11 | -0.17 | 0.00 |
| Hindu or Not | 0.78 | 0.77 | 0.75 | 0.77 | 0.06 | 0.00 |
| Degree of Cluster Rurality | 0.58 | 0.45 | 0.28 | 0.45 | 1.34 | 0.01 |

| Lesotho 2014 | | | | | | |
|---|---|---|---|---|---|---|
| Age | 28.96 | 32.66 | 34.41 | 32.40 | -0.47 | 0.02 |
| Body Mass Index | 20.85 | 21.36 | 22.15 | 21.49 | -0.41 | -0.04 |
| Wealth Index | 2.45 | 2.59 | 2.84 | 2.61 | -0.31 | -0.01 |
| Education Level | 1.07 | 1.17 | 1.32 | 1.18 | -0.36 | -0.03 |
| Currently Married or Not | 0.34 | 0.50 | 0.61 | 0.48 | -0.57 | 0.05 |
| Christian or Not | 0.89 | 0.90 | 0.92 | 0.90 | -0.10 | 0.02 |
| Degree of Cluster Rurality | 0.67 | 0.53 | 0.36 | 0.49 | 1.32 | 0.17 |

| Namibia 2013 | | | | | | |
|---|---|---|---|---|---|---|
| Age | 36.87 | 35.81 | 30.67 | 36.31 | 0.48 | -0.04 |

| | | | | | | |
|---|---|---|---|---|---|---|
| Body Mass Index | 21.63 | 21.39 | 20.46 | 21.60 | 0.31 | -0.06 |
| Wealth Index | 2.65 | 2.55 | 2.35 | 2.56 | 0.26 | -0.02 |
| Education Level | 1.09 | 1.15 | 1.41 | 1.13 | -0.41 | 0.02 |
| Currently Married or Not | 0.53 | 0.47 | 0.31 | 0.49 | 0.47 | -0.06 |
| Christian or Not | 0.80 | 0.81 | 0.86 | 0.78 | -0.18 | 0.06 |
| Degree of Cluster Rurality | 0.57 | 0.43 | 0.09 | 0.39 | 2.20 | 0.19 |

| Senegal 2010 | | | | | | |
|---|---|---|---|---|---|---|
| Age | 31.24 | 31.53 | 30.12 | 31.55 | 0.09 | 0.00 |
| Body Mass Index | 20.07 | 20.53 | 20.54 | 20.47 | -0.14 | 0.02 |
| Wealth Index | 1.75 | 2.00 | 2.39 | 2.00 | -0.63 | 0.01 |
| Education Level | 0.54 | 0.62 | 0.74 | 0.61 | -0.25 | 0.01 |
| Currently Married or Not | 0.54 | 0.54 | 0.48 | 0.54 | 0.12 | -0.01 |
| Christian or Not | 0.47 | 0.04 | 0.03 | 0.03 | 0.09 | 0.03 |
| Muslim or Not | 0.94 | 0.96 | 0.97 | 0.97 | -0.13 | -0.03 |
| Degree of Cluster Rurality | 0.70 | 0.53 | 0.33 | 0.50 | 1.54 | 0.15 |

| Uganda 2016 | | | | | | |
|---|---|---|---|---|---|---|
| Age | 30.73 | 28.99 | 28.17 | 29.15 | 0.23 | -0.01 |
| Body Mass Index | 20.60 | 20.84 | 21.17 | 20.90 | -0.21 | -0.02 |
| Wealth Index | 2.34 | 2.70 | 3.08 | 2.73 | -0.60 | -0.02 |
| Education Level | 1.24 | 1.35 | 1.46 | 1.38 | -0.32 | -0.04 |
| Currently Married or Not | 0.63 | 0.56 | 0.53 | 0.56 | 0.20 | 0.00 |
| Christian or Not | 0.90 | 0.88 | 0.86 | 0.88 | 0.13 | 0.00 |
| Degree of Cluster Rurality | 0.70 | 0.55 | 0.34 | 0.52 | 1.49 | 0.14 |

| Zimbabwe 2015 | | | | | | |
|---|---|---|---|---|---|---|
| Age | 29.52 | 29.01 | 28.03 | 29.05 | 0.14 | 0.00 |
| Body Mass Index | 20.90 | 20.90 | 21.04 | 20.90 | -0.05 | 0.00 |
| Wealth Index | 2.40 | 2.38 | 2.50 | 2.41 | -0.09 | -0.03 |
| Education Level | 1.57 | 1.60 | 1.73 | 1.63 | -0.28 | -0.05 |
| Currently Married or Not | 0.55 | 0.53 | 0.49 | 0.52 | 0.13 | 0.01 |
| Christian or Not | 0.69 | 0.70 | 0.74 | 0.69 | -0.11 | 0.00 |
| No Religion or Not | 0.25 | 0.25 | 0.22 | 0.25 | 0.06 | 0.00 |
| Degree of Cluster Rurality | 0.41 | 0.34 | 0.21 | 0.32 | 1.08 | 0.10 |

| Whole DHS Sample | | | | | | |
|---|---|---|---|---|---|---|
| | Agricultural Male Workers | | Better-off Controls | | Standardized Difference | |
| **Covariates** | Mean Before Matching | Mean After Matching | Mean Before Matching | Mean After Matching | Std Before Matching | Std After Matching |
| Age | 34.32 | 33.77 | 33.74 | 33.84 | 0.06 | -0.01 |
| Body Mass Index | 20.99 | 22.11 | 22.61 | 22.01 | -0.45 | 0.03 |
| Wealth Index | 2.38 | 2.96 | 3.32 | 2.95 | -0.75 | 0.01 |
| Education Level | 1.27 | 1.79 | 2.04 | 1.77 | -0.88 | 0.02 |
| Currently Married or Not | 0.74 | 0.72 | 0.72 | 0.73 | 0.05 | -0.03 |
| Christian or Not | 0.23 | 0.18 | 0.18 | 0.18 | 0.13 | 0.00 |
| Muslim or Not | 0.15 | 0.18 | 0.19 | 0.18 | -0.11 | 0.00 |
| Hindu or Not | 0.56 | 0.58 | 0.58 | 0.58 | -0.03 | 0.00 |
| No Religion or Not | 0.01 | 0.01 | 0.01 | 0.01 | 0.02 | 0.01 |
| Degree of Cluster Rurality | 0.29 | 0.41 | 0.29 | 0.41 | 1.38 | 0.03 |

| Whole DHS Sample | | | | | | |
|---|---|---|---|---|---|---|
| | Agricultural Male Workers | | Worse-off Controls | | Standardized Difference | |
| Age | 34.32 | 31.81 | 28.16 | 31.89 | 0.56 | -0.01 |
| Body Mass Index | 20.99 | 21.11 | 20.88 | 21.14 | 0.03 | -0.01 |
| Wealth Index | 2.38 | 2.50 | 2.65 | 2.50 | -0.22 | 0.00 |
| Education Level | 1.27 | 1.49 | 1.67 | 1.49 | -0.46 | 0.00 |
| Currently Married or Not | 0.74 | 0.64 | 0.49 | 0.65 | 0.54 | -0.01 |
| Christian or Not | 0.23 | 0.19 | 0.18 | 0.20 | 0.13 | 0.00 |
| Muslim or Not | 0.15 | 0.14 | 0.14 | 0.14 | 0.01 | 0.00 |
| Hindu or Not | 0.56 | 0.60 | 0.61 | 0.59 | -0.10 | 0.00 |
| No Religion or Not | 0.01 | 0.02 | 0.02 | 0.02 | -0.07 | 0.00 |
| Degree of Cluster Rurality | 0.62 | 0.45 | 0.28 | 0.45 | 1.44 | 0.01 |

| Whole DHS Sample | | | | | | |
|---|---|---|---|---|---|---|
| | Worse-off Controls | | Better-off Controls | | Standardized Difference | |
| Age | 28.16 | 32.54 | 33.74 | 32.56 | -0.54 | 0.00 |
| Body Mass Index | 20.88 | 22.23 | 22.61 | 22.18 | -0.47 | 0.02 |
| Wealth Index | 2.65 | 3.09 | 3.18 | 3.09 | -0.53 | 0.00 |
| Education Level | 1.67 | 1.86 | 2.04 | 1.87 | -0.45 | -0.01 |
| Currently Married or Not | 0.49 | 0.66 | 0.72 | 0.66 | -0.49 | 0.00 |
| Christian or Not | 0.18 | 0.18 | 0.18 | 0.18 | -0.01 | 0.00 |
| Muslim or Not | 0.14 | 0.18 | 0.19 | 0.19 | -0.12 | -0.01 |
| Hindu or Not | 0.61 | 0.58 | 0.58 | 0.58 | 0.07 | 0.00 |
| No Religion or Not | 0.02 | 0.01 | 0.01 | 0.01 | 0.09 | 0.02 |
| Degree of Cluster Rurality | 0.28 | 0.29 | 0.29 | 0.29 | -0.05 | 0.01 |

#### Section 5. One-Parameter Sensitivity analysis

Part I. Result of one-parameter sensitivity analysis for the whole sample and each DHS sample with maximum p-value < 0.05 when Γ = 1 (agricultural workers vs. all controls)

Whole DHS Sample

| Γ | P-value lower bound for one-sided test | P-value upper bound for one-sided test |
|---|---|---|
| 1.00 | $< 10^{-8}$ | $< 10^{-8}$ |
| 1.06 | $< 10^{-12}$ | 0.008 |
| 1.07 | $< 10^{-12}$ | 0.029 |
| 1.08 | $< 10^{-12}$ | 0.085 |

Ethiopia 2016

| Γ | P-value lower bound for one-sided test | P-value upper bound for one-sided test |
|---|---|---|
| 1.00 | 0.008 | 0.008 |
| 1.04 | 0.001 | 0.035 |
| 1.05 | 0.001 | 0.047 |
| 1.06 | $< 10^{-4}$ | 0.063 |

India 2015

| Γ | P-value lower bound for one-sided test | P-value upper bound for one-sided test |
|---|---|---|
| 1.00 | $< 10^{-4}$ | $< 10^{-4}$ |
| 1.03 | $< 10^{-7}$ | 0.006 |
| 1.04 | $< 10^{-8}$ | 0.021 |
| 1.05 | $< 10^{-10}$ | 0.058 |

Lesotho 2014

| Γ | P-value lower bound for one-sided test | P-value upper bound for one-sided test |
|---|---|---|
| 1.00 | 0.012 | 0.012 |
| 1.08 | 0.003 | 0.041 |
| 1.09 | 0.002 | 0.047 |
| 1.10 | 0.002 | 0.053 |

Senegal 2010

| Γ | P-value lower bound for one-sided test | P-value upper bound for one-sided test |
|---|---|---|
| 1.00 | 0.029 | 0.029 |
| 1.01 | 0.023 | 0.036 |
| 1.02 | 0.018 | 0.044 |
| 1.03 | 0.014 | 0.053 |

Part II. Result of sensitivity analysis for agricultural male workers vs all controls, agricultural male workers vs better-off controls, agricultural male workers vs worse-off controls, and worse-off control vs. better-off control for the whole sample.

Agricultural Workers vs. All Controls

| Γ | P-value lower bound for one-sided test | P-value upper bound for one-sided test |
|---|---|---|
| 1.00 | $< 10^{-8}$ | $< 10^{-8}$ |
| 1.06 | $< 10^{-12}$ | 0.008 |
| 1.07 | $< 10^{-12}$ | 0.029 |
| 1.08 | $< 10^{-12}$ | 0.085 |

Agricultural Workers vs. Better-off Controls

| Γ | P-value lower bound for one-sided test | P-value upper bound for one-sided test |
|---|---|---|
| 1.00 | $< 10^{-10}$ | $< 10^{-10}$ |
| 1.14 | $< 10^{-12}$ | 0.017 |
| 1.15 | $< 10^{-12}$ | 0.034 |
| 1.16 | $< 10^{-12}$ | 0.062 |

Agricultural Workers vs. Worse-off Controls

| Γ | P-value lower bound for one-sided test | P-value upper bound for one-sided test |
|---|---|---|
| 1.00 | $< 10^{-7}$ | $< 10^{-7}$ |
| 1.06 | $< 10^{-12}$ | 0.011 |
| 1.07 | $< 10^{-12}$ | 0.037 |
| 1.08 | $< 10^{-12}$ | 0.098 |

Worse-off Controls vs. Better-off Controls

| Γ | P-value lower bound for one-sided test | P-value upper bound for one-sided test |
|---|---|---|
| 1.00 | $< 10^{-3}$ | $< 10^{-3}$ |
| 1.04 | $< 10^{-6}$ | 0.020 |
| 1.05 | $< 10^{-7}$ | 0.045 |
| 1.06 | $< 10^{-8}$ | 0.090 |
